# The application of Hybrid deep learning Approach to evaluate chest ray images for the diagnosis of pneumonia in children

**DOI:** 10.1101/2020.12.03.20243550

**Authors:** Mohammad Ali Abbasa, Syed Usama Khalid Bukhari, Syed Khuzaima Arssalan Bokhari, manal niazi

**Affiliations:** Department of Computer Science, University of Lahore, Islamabad, Pakistan; Pediatric medicine, Doctors hospital, Lahore, Pakistan; Radiology department, Islamabad medical and dental college, Islamabad, Pakistan

**Keywords:** Deep Learning, Chest X-Ray, Pneumonia

## Abstract

**Background:** Pneumonia is a leading cause of morbidity and mortality worldwide, particularly among the developing nations. Pneumonia is the most common cause of death in children due to infectious etiology. Early and accurate Pneumonia diagnosis could play a vital role in reducing morbidity and mortality associated with this ailment. In this regard, the application of a new hybrid machine learning vision-based model may be a useful adjunct tool that can predict Pneumonia from chest X-ray (CXR) images.

**Aim & Objective:** we aimed to assess the diagnostic accuracy of hybrid machine learning vision-based model for the diagnosis of Pneumonia by evaluating chest X-ray (CXR) images

**Materials & Methods:** A total of five thousand eight hundred and fifty-six digital X-ray images of children from ages one to five were obtained from the Chest X-Ray Pneumonia dataset using the Kaggle site. The dataset contains fifteen hundred and eighty-three digital X-ray images categorized as normal, where four thousand two hundred and seventy-three digital X-ray images are categorized as Pneumonia by an expert clinician. In this research project, a new hybrid machine learning vision-based model has been evaluated that can predict Pneumonia from chest X-ray (CXR) images. The proposed model is a hybrid of convolutional neural network and tree base algorithms (random forest and light gradient boosting machine). In this study, a hybrid architecture with four variations and two variations of ResNet architecture are employed, and a comparison is made between them.

**Results:** In the present study, the analysis of digital X-ray images by four variations of hybrid architecture RN-18 RF, RN-18 LGBM, RN-34 RF, and RN-34 LGBM, along with two variations of ResNet architecture, ResNet-18 and ResNet-30 have revealed the diagnostic accuracy of 97.78%, 96.42%, 97.1%,96.59%, 95.05%, and 95.05%, respectively.

**Discussion:** The analysis of the present study results revealed more than 95% diagnostic accuracy for the diagnosis of Pneumonia by evaluating chest x-ray images of children with the help of four variations of hybrid architectures and two variations of ResNet architectures. Our findings are in accordance with the other published study in which the author used the deep learning algorithm Chex-Net with 121 layers.

**Conclusion:** The hybrid machine learning vision-based model is a useful tool for the assessment of chest x rays of children for the diagnosis of Pneumonia.

## 1 Introduction

The lungs are exposed to several microbial organisms that are present in the inhaled air. The microbial infection of the lungs is the primary etiological factor of pneumonia [1]. Bacteria are the primary pathogenic organisms that cause pneumonia. The other possible organisms include viruses and fungi. The microbial infection of the lung causes damage to the respiratory epithelium and stimulates an inflammatory response. The involvement of alveoli causes a problem with the oxygen exchange that can lead to death if not adequately treated at an early stage. Due to pneumonia, childhood mortality is approximately one million deaths per year of children less than five years of age [2]. A rapid and accurate diagnosis of pneumonia is vital for the appropriate management of this disease. The clinical features and radiological findings, particularly on chest X-ray, have paramount importance in diagnosing pneumonia. The interpretation of radiological features on the chest x-ray requires the opinion of an expert radiologist.

One of the critical limitations in the radiological interpretation of chest x-rays of children include the scarcity of the twenty-four hours availability of radiologist in many health centers of the developing nations. The other limitation of chest x-ray evaluation is the variation in inter-observer interpretation [3,4]. In this regard, an intelligent automated system that can diagnose pneumonia from chest x-ray images could significantly help manage pneumonia cases, particularly in the emergency situations and the remote and far-flung areas where there is a lack of round-the-clock availability of the radiologists.

## 2 Purposed Methodology

Convolutional Neural Networks (CNN) are used widely for image analysis tasks, especially image classification. CNN architectures can easily be divided into two parts; part I contains the convolution and pooling layers and used for feature extraction purposes. Where part II contains the dense layers, and this part is used for classification purposes.

ResNet architecture gain popularity due to its architecture consisting of identity maps and residual function, which resists falling in saddle points. Similarly, tree-based algorithms such as Random Forest and Gradient Boosting are popular for classification jobs.

In this paper, we have introduced a hybrid architecture that uses ResNet-18 and ResNet-34 feature extraction ability, and combine it with tree base algorithm Random-Forest and Light-GBM for classification purposes.

Four variations of hybrid architecture are developed for this study.

1. RN-18 RF; ResNet-18 (feature extraction) + Random Forest (classification).
2. RN-18 LGBM; ResNet-18 (feature extraction) + Light GBM (classification).
3. RN-34 RF; ResNet-34 (feature extraction) + Random Forest (classification).
4. RN-34 LGBM; ResNet-34 (feature extraction) + Light GBM (classification).

Algorithm-1 and algorithm-2 presents the feature extraction and hybrid model steps. Figure-1 shows the working of the system in block format.

### Algorithm 1: Feature Extractionn Code

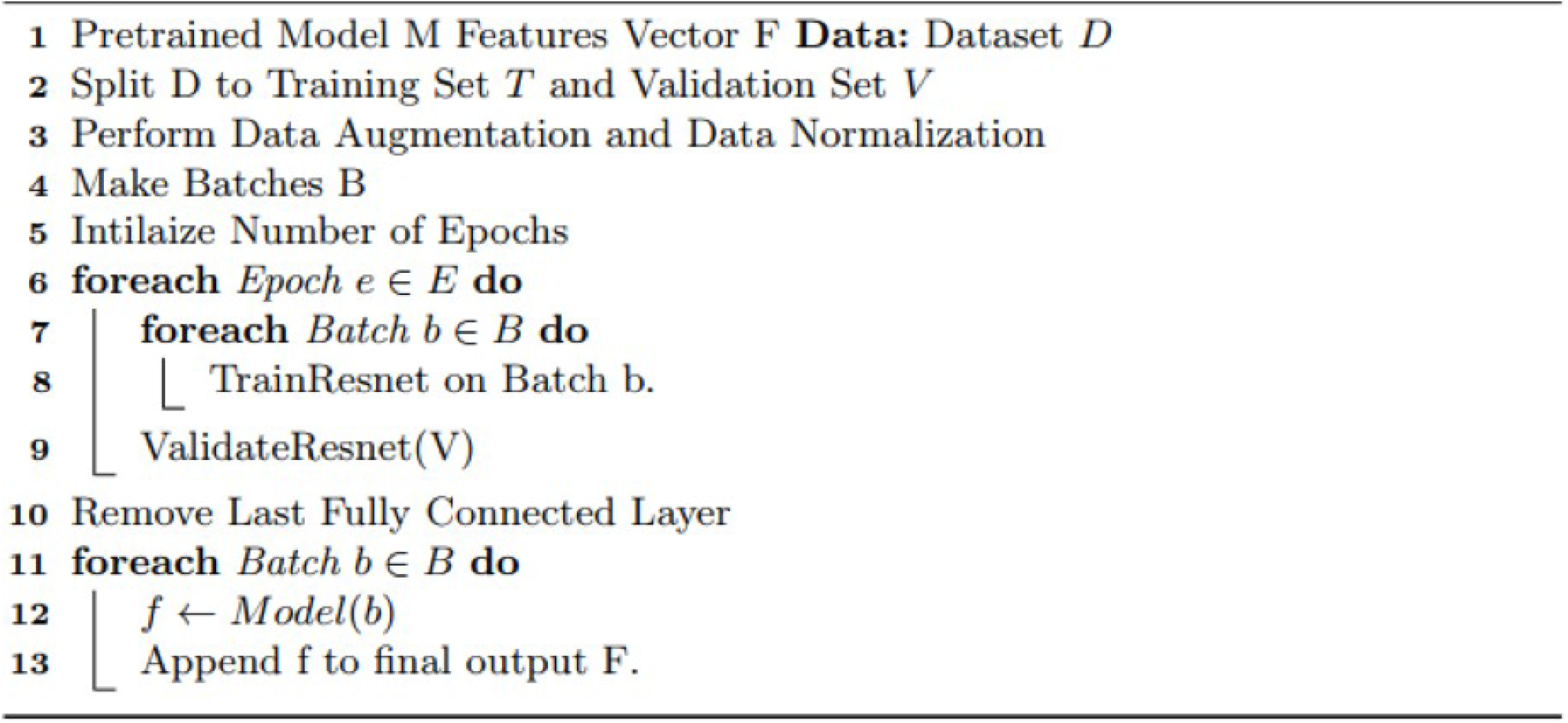

### Algorithm 2: Trainn Hybrid Models

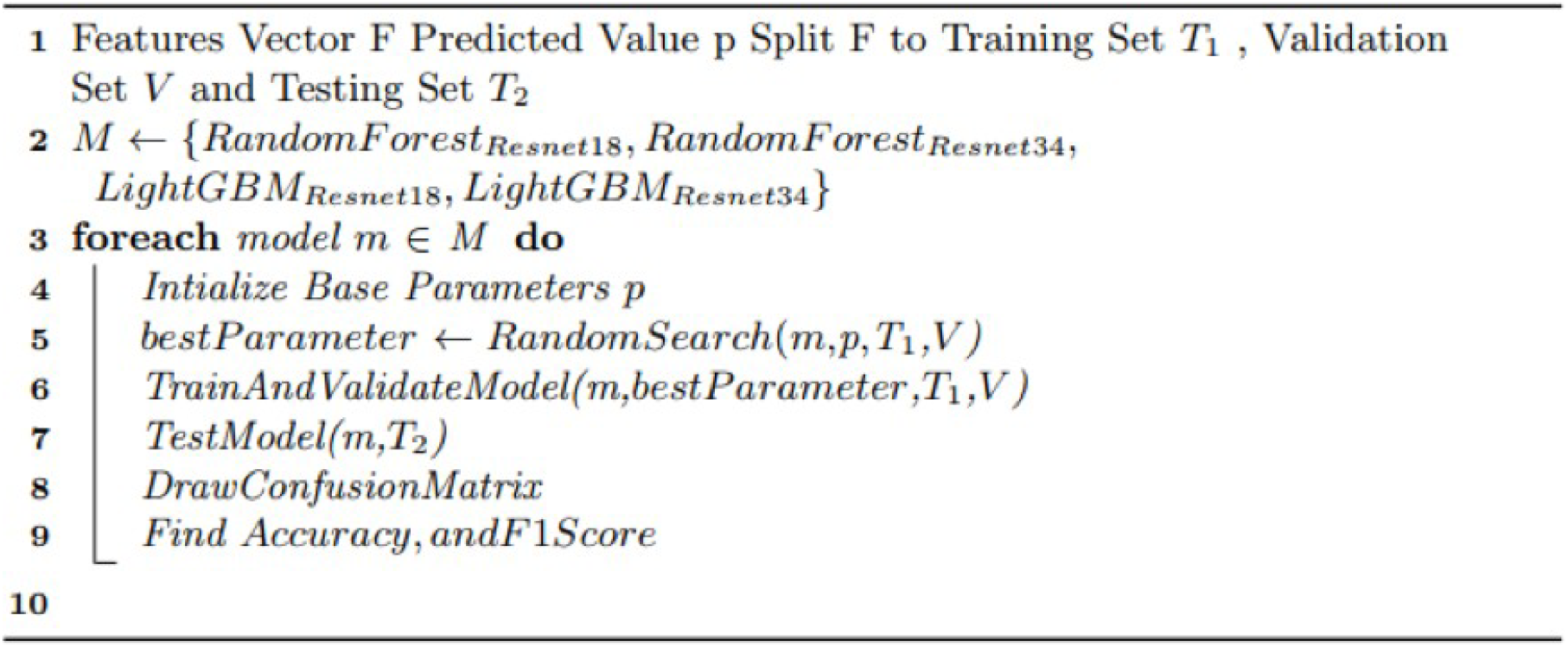

**Fig 1.**
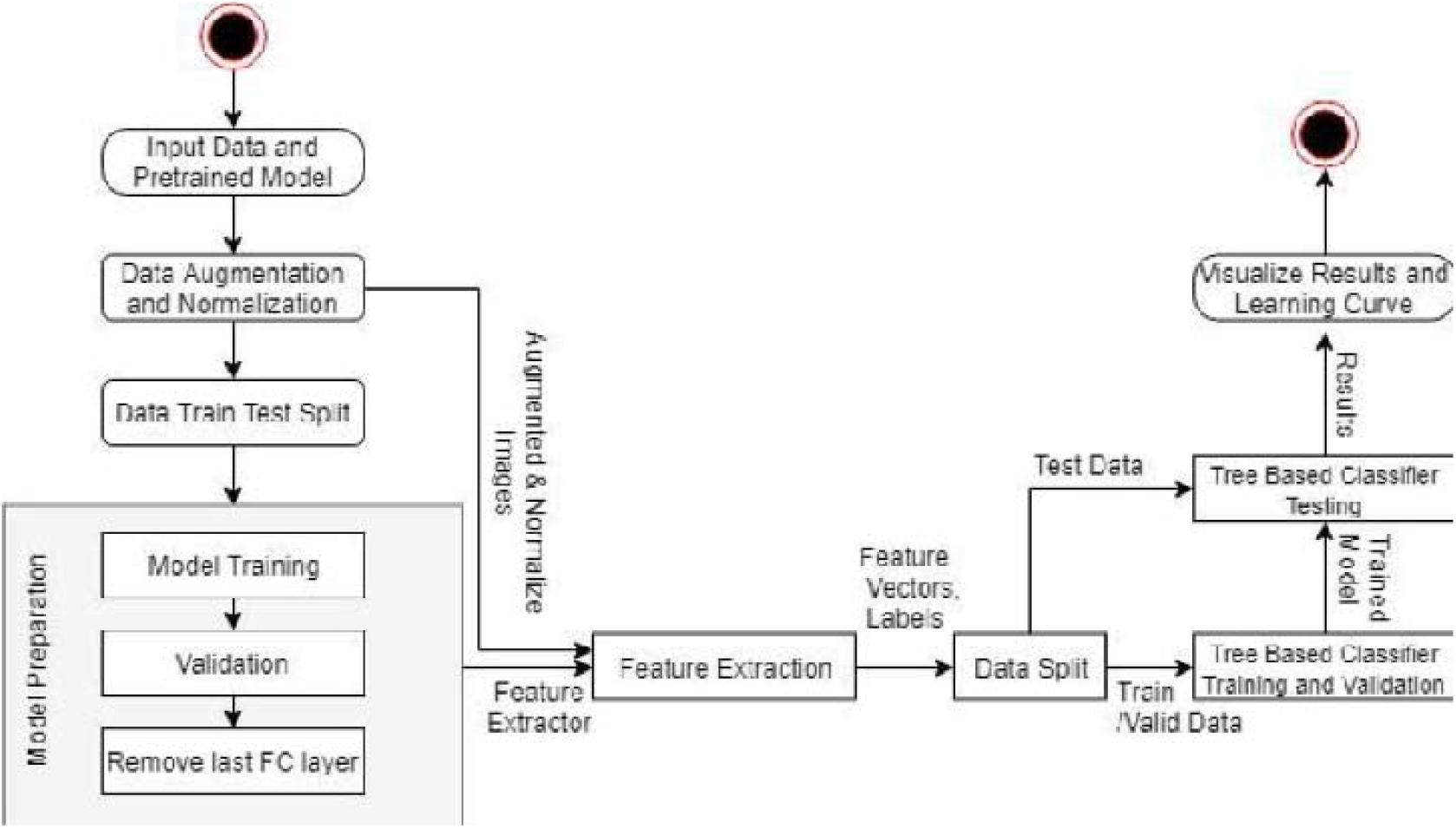
Flow chart of Hybrid Architecture.

## 3 Materials & Methods

A total of five thousand eight hundred and fifty-six digital (5856) X-ray images of children from ages one to five were obtained from the Chest X-Ray Pneumonia dataset [5] using the Kaggle site. The dataset contains fifteen hundred and eighty-three digital (1583) X-ray images categorized as normal, where four thousand two hundred and seventy-three digital (4373) X-ray images are categorized as Pneumonia by an expert physician. For accuracy, the expert clinician rechecked all the labeled data.

The data is divided into three sets (Train, Test, and Valid). The train set contained 70% of total data, four thousand and ninety-nine (4099) X-ray images of both normal and Pneumonia categories. The valid set contained 20% of total data, eleven hundred and seventy-one (1171) X-ray images of both categories, where the Test set contained 10% of the total data, five hundred and six (586) X-ray images. One hundred and fifty-eight (158) images of the normal category, and four hundred and twenty-eight (428) images of the pneumonia category.

Four variations of proposed hybrid architectures RN-18 RF, RN-18 LGBM, RN-34 RF, and RN-34 LGBM, two variations of ResNet, ResNet-18, and ResNet-34, are used analyze the dataset. For the training, Fast AI API’s pre-trained ResNet models (ResNet-18 and ResNet-34) on image net and standard data augmentation parameters are used, with a learning rate of 0.0006, momentum 0.3, and weight decay of 0.1 for 20 epochs. These newly trained ResNet models are then used to generate the results on Test sets and be used for feature extraction for our hybrid architectures. This procedure is followed so that the convolution part of CNN remains similar (same hyperparameters and training) for standard ResNet architecture and proposed hybrid architectures. The extracted features from both trained ResNet-18 and ResNet-34 are stored in two comma-separated value (CSV) files along with labels. Random forest and Light BGM tree classifiers (of ski learn library) are trained on these CSV files. After training for these algorithms, Test data (from Kaggle) is used to generate the final results.

## 4 Results

**Table 1.** Confusion Matrix of Test Datasets.

| Architectures | Test Data Set |  |  |  | Architectures | Test Data Set |  |  |  |
| --- | --- | --- | --- | --- | --- | --- | --- | --- | --- |
| Resnet 18 | Actual | Normal | 142 | 16 | Resnet 34 | Actual | Normal | 140 | 18 |
|  |  | Pneumonia | 13 | 415 |  |  | Pneumonia | 11 | 417 |
|  |  |  | Normal | Pneumonia |  |  |  | Normal | Pneumonia |
|  |  | Predicted |  |  |  |  | Predicted |  |  |
| RN-18 RF | Actual | Normal | 149 | 9 | RN-34 RF | Actual | Normal | 144 | 14 |
|  |  | Pneumonia | 4 | 424 |  |  | Pneumonia | 3 | 425 |
|  |  |  | Normal | Pneumonia |  |  |  | Normal | Pneumonia |
|  |  | Predicted |  |  |  |  | Predicted |  |  |
| RN-18 LGBM | Actual | Normal | 146 | 12 | RN-34 LGBM | Actual | Normal | 144 | 14 |
|  |  | Pneumonia | 9 | 419 |  |  | Pneumonia | 6 | 422 |
|  |  |  | Normal | Pneumonia |  |  |  | Normal | Pneumonia |
|  |  | Predicted |  |  |  |  | Predicted |  |  |

**Table 2.** Detail Results of Test Dataset.

| ARCHITECTURES | SENSITIVITY | SPECIFICITY | F-1 SCORE | ACCURACY |
| --- | --- | --- | --- | --- |
| **RESNET-18** | 0.916 | 0.962 | 0.907 | 95.05% |
| **RN-18 RF** | 0.973 | 0.979 | 0.958 | 97.78 % |
| **RN-18 LGBM** | 0.941 | 0.972 | 0.932 | 96.42 % |
| **RESNET-34** | 0.927 | 0.958 | 0.906 | 95.05% |
| **RN-34 RF** | 0.979 | 0.968 | 0.944 | 97.1 % |
| **RN-34 LGBM** | 0.960 | 0.967 | 0.935 | 96.59 % |

## 5 Discussion

In the present study, the application of four variations of hybrid architectures and two variations of ResNet architectures for the evaluation of chest x-ray images revealed more than 95% diagnostic accuracy for the diagnosis of pneumonia in children.

The different techniques had been used to evaluate the chest X-rays, but most of these techniques are not very applicable. The use of machine learning helped improve the algorithms, but the accuracy of the systems was pretty low [5,6]. In 2007 onward, deep learning and Convolution Neural Network (CNN) made its impact, and things started to go in a positive direction [7]. Deep learning algorithms have a built-in flaw that they need a big data set to train themselves. Hence many institutes and researchers started to develop the data sets. In 2017 Wang et al. released a data set of chest X-ray images named ChestXray8, comprising 108,948 frontal view X-ray images with eight diseases of 32,717 unique patients [8]. This is also known as the National Institute of Health (NIH) CXR images dataset. This data set comprises 100,000 frontal-view X-ray images with 14 diseases [9]. In his paper, Rajpurker et al. presented a new deep learning algorithm, CheXNet, with 121 layers, that shows its results were as good as an expert radiologist. The F1-score of algorithms was almost 0.71, equal to a human radiologist average score [9]. Ivo M. Baltruschat et al., in 2019, presented a comparison of different deep learning models for X-ray image classification problems [10]. In these models, Resnet-34 and Resnet-50 were trained using transfer learning.

In comparison, the highest accuracy that has been achieved was 76 % for the pneumonia detection by any model. Okeke Stephen et al. developed a convolutional neural network model from scratch to perform binary CXR images classification as pneumonia or healthy persons [11]. The model was tested on different test sizes, from 100 images to 400 images. The model achieved an average accuracy of 94%. There are several research methods in research that provide accuracy between 70 to 94% for the pneumonia classification problem. As this problem is linked with human lives hence increasing inaccuracy in detection is essential.

Hybrid approaches to merge the CNN and other ML algorithms are becoming popular in literature as, in different cases, they outperformed existing state of the art. Xiao-XiaoNiu et al. developed a Hybrid Algorithm by Combination of CNN and Support Vector Machine (SVM) and achieved an accuracy of about 99% for digit recognition [7]. In his paper, Ben Athiwaratkun et al. presented different hybrid algorithms and shows they performed better [2]. Shaoqing Ren et al. used Faster R-CNN for feature extraction by removing a multi-perceptron layer of CNN, and features are passed to Random Forest, and it provided better results than CNN [9].

This paper presents a new hybrid approach to do the binary classification between pneumonia and healthy patients using CRX images. To develop the proposed hybrid method, we used the Resnet [12] with random forest [13] and Microsoft’s light Gradient Boosting Machines (BGM) algorithm [14]. The results show that the accuracy of the hybrid method is much better than any other available method.

## 6 Conclusion

The findings of this study raise the possibility of using a hybrid machine learning vision-based model as a tool for assessing chest x rays of children for the diagnosis of pneumonia. The application of convolutional neural networks for the radiological diagnosis of pneumonia may assist the clinician in emergency situations, particularly in the regions without radiologists’ 24-hour availability. It may also assist the radiologist and may reduce the stress and workload on them. The availability of datasets of radiology images labeled by an expert radiologist may be a useful learning resource for the trainees.

## Data Availability

https://stanfordmlgroup.github.io/projects/chexnet/

